# Point-of-care digital cytology with artificial intelligence for cervical cancer screening at a peripheral clinic in Kenya

**DOI:** 10.1101/2020.08.12.20172346

**Authors:** Oscar Holmström, Nina Linder, Harrison Kaingu, Ngali Mbuuko, Jumaa Mbete, Felix Kinyua, Sara Törnquist, Martin Muinde, Leena Krogerus, Mikael Lundin, Vinod Diwan, Johan Lundin

**Affiliations:** Institute for Molecular Medicine Finland (FIMM), Tukholmankatu 8, University of Helsinki, Helsinki, Finland; Department of Women’s and Children’s Health, International Maternal and Child Health, Uppsala University, SE-751 85 Uppsala, Sweden; Kinondo Kwetu Health Services Clinic, Kinondo, Kwale County, Kenya; Department of Global Public Health, Karolinska Institutet, 171 77 Stockholm, Sweden; Helsinki University Hospital and HUSLAB Pathology Laboratory, Tukholmankatu 8, Helsinki, Finland

**Author notes:** **Corresponding author:** Johan Lundin, Institute for Molecular Medicine Finland (FIMM), P.O. Box 20, FI-00014, University of Helsinki, Finland.

## Abstract

Cervical cancer is highly preventable but remains a common and deadly cancer in areas without screening programmes. Pap smear analysis is the most commonly used screening method but is labour-intensive, subjective and requires access to medical experts. We developed a diagnostic system in which microscopy samples are digitized at the point-of-care (POC) and analysed by a cloud-based deep-learning system (DLS) and evaluated the system for the detection of cervical cell atypia in Pap smears at a peripheral clinic in Kenya. A total of 740 conventional Pap smears were collected, digitized with a portable slide scanner and uploaded over mobile networks to a cloud server for training and validation of the system. In total, 16,133 manually-annotated image regions where used for training of the DLS. The DLS achieved a high average sensitivity (97.85%; 95% confidence interval (CI) 83.95—99.75%) and area under the curve (AUCs) (0.95) for the detection of cervical-cellular atypia, compared to the pathologist assessment of digital and physical slides. Specificity was higher for high-grade atypia (95.9%; 95% CI 94.9—97.6%) than for low-grade atypia (84.2%; 95% CI 79.9—87.9%). Negative predictive values were high (99.3-100%), and no samples classified as high grade by manual sample analysis had false-negative assessments by the DLS. The study shows that advanced digital microscopy diagnostics supported by machine learning algorithms is implementable in rural, resource-constrained areas, and can achieve a diagnostic accuracy close to the level of highly trained experts.

**Summary box:** *What is already known?:*

• Cervical cancer can be prevented with Pap smear screening, but manual sample analysis is labor-intensive, subjective and not widely-available in regions with the highest disease prevalence
• Novel digital methods, such as image-based artificial intelligence (AI), show promise for facilitated analysis of microscopy samples
• Digital methods are typically limited to high-end laboratories, due to the requirements for advanced equipment and supportive digital infrastructure

*What are the new findings?:*

• A point-of-care diagnostic system where samples are digitized with a portable slide scanner and analyzed using a cloud-based AI model can be implemented in rural settings and utilized to automatically interpret Pap smears and identify potentially precancerous samples with similar accuracy as a pathologist specialized in reading Pap smears.

*What do the new findings imply?:*

• The results demonstrate how advanced digital methods, such as AI-based digital microscopy, can be implemented in rural, resource-limited areas, and used for analysis of microscopy samples, such as Pap smears.
• This technology shows promise as a novel method for digital microscopy diagnostics, which can be implemented in rural settings, and could be of particular value in areas lacking cytotechnicians and pathologists.

## Introduction

Inadequate access to microscopy diagnostics affects over a billion people in low-resource areas and causes the underdiagnosis of a number of common and treatable conditions.^1^ Although significant advances have been made in technologies for microscopy diagnostics at the point-of-care (POC), their clinical implementation has been slow.^2^ In this article, we propose a novel digital diagnostics system in which microscopy slides are digitized at the POC and uploaded using local data networks for analysis with an artificial intelligence (AI) model based on deep convolutional neural networks. We implemented the system in the setting of a peripheral clinic in Kenya and evaluated its use for the analysis of cervical smears (Papanicolaou (Pap) smears). Cervical cancer is a significant health problem in areas without screening programmes, where it remains among the most common and deadliest cancers.^3^ During the next decade, the disease incidence is predicted to increase, and the yearly mortality is expected to double, with the largest burden of disease occurring in sub-Saharan Africa.^4^ As the causative agent of cervical cancer is human papillomavirus (HPV),^5^ it has been possible to develop several efficient prevention methods.^3^ Theoretically, HPV vaccines have the potential to eradicate the disease, but as the full benefits of even the most efficient vaccination programmes will take decades to realize, millions of women who are already infected with HPV remain at risk.^6^ Therefore, screening tests remain essential,^7^ and innovative POC diagnostic solutions are needed.^8^ Conventional cytology screening (Pap smear analysis) can drastically reduce the incidence and mortality of cervical cancer when it is adequately implemented, but the manual analysis of samples is labour intensive,^9^ prone to variations in sensitivity and reproducibility, and requires medical experts to analyse the samples,^10,11^ thus making it hard to implement in resource-limited settings.^12^ Molecular techniques that detect the nucleic acid of oncogenic HPV have high sensitivity and reproducibility but are limited by the requirement for validated laboratory assays and a relatively low specificity, especially in high-risk populations, as most HPV infections are transient.^13,14^ In high-resource areas, both molecular- and cytology-based screening are commonly used and are often combined (‘cotesting’) to improve the diagnostic accuracy.^15,16^ Digital methods have been proposed to facilitate the visual analysis of Pap smears, but the development of fully automated systems has been challenging.^17,18^ Although semi-automatic systems for Pap smear screening have been developed,^19^ they are limited by the need for bulky, expensive laboratory equipment^20,21^ and are not suitable for usage in POC or rural settings. Traditional image analysis methods have relied on the identification of pre-defined cellular features in digitized slides (by methods such as the segmentation of the nucleus and cytoplasm of individual cells), rendering them vulnerable to sample quality problems, such as debris and overlapping cells.^19,22^ Recently, deep learning-based medical AI algorithms have provided efficient tools for medical image analysis applications, with levels of performance that have even surpassed human experts in certain tasks.^23-26^ However, deep learning algorithms have been mainly studied for the analysis of cervical cytology smears using cropped images from digital samples with a limited number of cells that have been digitized with high-end equipment; however, to our knowledge, no research has been conducted on the analysis of whole slides prepared in rural clinical environments.^26-29^ Thus, the technology has so far not been applied in locations without access to well-equipped laboratories, where the need for improved diagnostics is highest.^28^

In this study, we developed and implemented a novel POC digital diagnostic system at a rural clinic in Kenya, a country where cervical cancer is the leading cause of female cancer-related death.^30^ Pap smears collected at the clinic were digitized with a portable slide scanner, and whole-slide images were uploaded to a cloud server using the local mobile data network for analysis by a deep learning system (DLS). We measured the diagnostic accuracy for the detection of common forms of cervical squamous cell atypia with the DLS and validated the results by comparing them with the visual assessment of both digital and physical slides by independent pathologists.

## Methods

### Study design, patient cohort and collection of samples

This study is a diagnostic accuracy study which is reported in accordance with the STARD-guidelines (Supplementary Figure 1). The research site for this study was a local clinic (Kinondo Kwetu Health Services Clinic, Kinondo, Kwale County) in Kenya (Figure 1). Pap smears (n = 740) for this study were acquired from women in a regional HIV-control programme in Kwale County. Samples were collected from volunteering patients who fulfilled the inclusion criteria: non-pregnant women, aged between 18 and 64 years (mean: 41.76), confirmed HIV-positivity (mean year of diagnosis 2012), and signed informed consent acquired. In this cohort, a minority (2.6%) reported that they had previously participated in Pap smear screening. The reported mean number of children per patient was 3.39 (SD 2.16), a minority of patients were smokers (1.8%), 28.7% were postmenopausal and no patients were currently receiving hormonal replacement therapy (Supplementary Table 1). Eligible patients were assigned a study number, after which Pap smears were obtained from the patients using a cervical broom sampling kit (Touchfree Cytopak, AS Diagnostics & Disposables, Chennai, India) by trained nurses.

**Figure 1.**
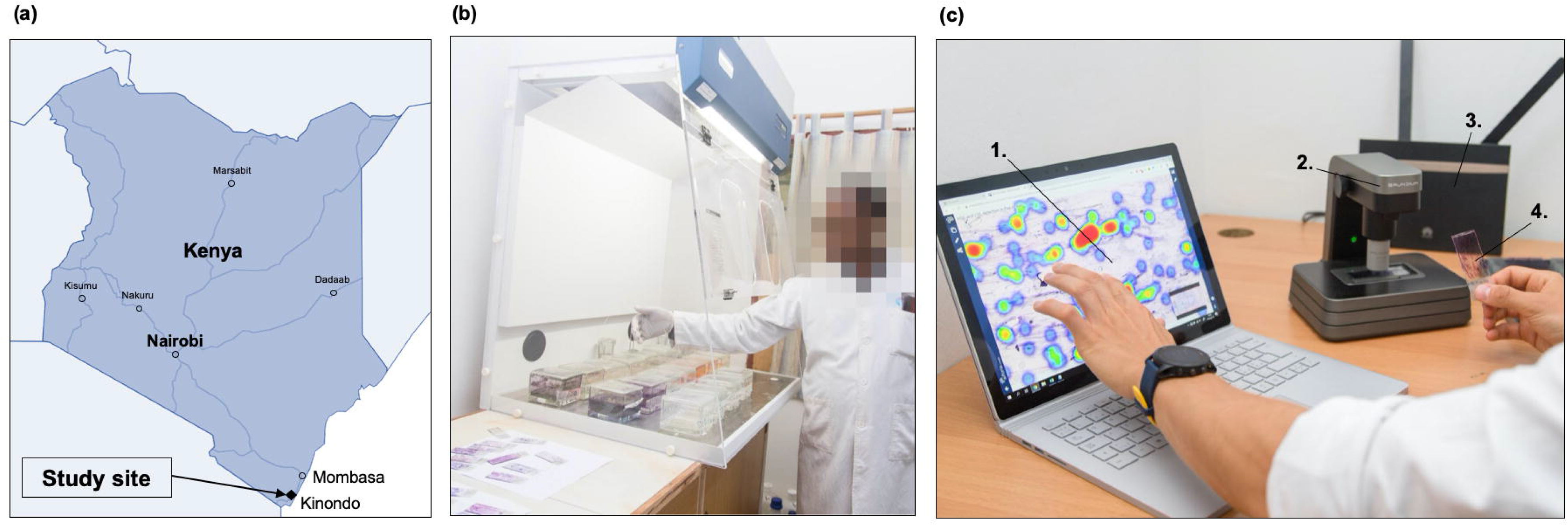
Practical aspects of the study methodology. (a) Location of the study site in Kenya. (b) Slide staining bench and hood. (c) Slide digitization equipment: (1) laptop computer with access to the slide-management platform; (2) slide scanner; (3) mobile-network router; and (4) Pap-smear microscopy slide.

The cervical sample was applied to a clean frosted glass slide, fixated using the provided fixative solution and allowed to air-dry at room temperature. Following sample acquisition, the slides were fixed in 95% ethyl alcohol for 15 minutes at room temperature after which staining was performed using glassware equipment placed in a fume hood, with the Papanicolaou staining method.^31^ Laboratory reagents for the staining process were acquired from Cytolab Kenya (Cytolab Enterprises, Nairobi, Kenya). For the staining process, the slides were first immersed in 70% and 50% ethyl alcohol, consequently, and rinsed with distilled water. Following this, the slides were stained using Meyer’s haematoxylin, after which they were washed in distilled water and 95% ethyl alcohol. The first counterstain was performed using the OG-6 staining solution, after which the slides were again washed in 95% ethyl alcohol. The second counterstain was performed using the EA-65 staining solution, after which slides were washed in 95% ethyl alcohol and dehydrated using absolute ethanol. Sample clearance was after this carried out with rectified Xylene. After the staining procedure, the slides were covered with coverslips using the diethyl propene xylene (DPX) mounting medium, and air-dried at room temperature in a level position overnight prior to analysis. Stained slides were examined using a light microscope to assess quality before further analyses and stored in slide boxes before digitization and transportation to the pathology laboratory (Coast Provincial General Hospital, Mombasa, Kenya). Prior to this, slide labels were counter-checked against the patient records and the numbers recorded on the report forms which were sent with the slide boxes to the pathologist laboratory. Patients with slides which were classified as inadequate for analysis by the pathologist, were contacted and offered to provide a repeat sample for analysis. Clinical patient records used in the study were stored in digital format using the secured and password-protected web-based data-collection software REDCap (Research Electronic Data Capture, Vanderbilt University, Nashville, TN, USA), running on a password-protected, encrypted local server in a locked room. Paper forms with patient data were stored in locked cabinets in a locked room at the clinic, accessible only to study personnel. Both digitized and physical slides were pseudonymized using study numbers and no personal identifiers were uploaded to the image-management platform. Prior to study participation, eligible patients were given information in both written and oral form about the purpose of the study and the testing procedure. Information was provided in English and Swahili before signed consent from patients was obtained. Patients were compensated for travel expenses to the sample-acquisition site and informed of the test results, but were not offered other monetary compensation for study participation. In cases of abnormal Pap smears, treatment expenses were covered by study funding, and treatment was arranged by a gynaecologist (JM) in accordance with national guidelines.^32^

### Digitization of slides at the research site

Following the acquisition and staining of samples, Pap smears were digitized with a portable whole-slide microscope scanner (Grundium Ocus, Grundium, Tampere, Finland) (Figure 1). The device features an 18-megapixel image sensor with a 20× objective (NA 0.40) and a pixel size of 0.48 μm. The microscope was connected to a laptop computer over a wireless local area network connection and operated via a browser interface. The coarse focus for the scanner was adjusted manually, after which the built-in autofocus routine was utilized for fine focus. Image files were saved on the local computer in Tagged Image File Format and converted to a wavelet file format (Enhanced Compressed Wavelet, Hexagon Geospatial, Madison, AL, USA) using a compression ratio (1:16) that was previously shown to preserve sufficient detail to not significantly alter the image-analysis results,^33^ before uploading to the image-management and machine-learning platform (Aiforia Cloud and Aiforia Create, Aiforia Technologies, Helsinki, Finland). On the slide-management platform, digitized slides were stored as JPG-compressed tile-maps with a pyramid-structure of zoom levels (70% JPG-quality). Uploading of slides was performed primarily via a 3G/4G mobile-network router (Huawei B525S, Huawei Technologies, Shenzhen, China) operating on the local mobile network (Safaricom, Nairobi, Kenya), with a subset of slides uploaded via the in-house ADSL connection. The compressed size of the digitized slides ranged from 0.2 Mb to 0.8 Gb, resulting in a turnaround time for sample uploading of ~10-40 min over the mobile network (upload speed 5-8 Mbps) or ADSL connection (upload speed 5-10Mbps). Access to the image server for remote slide viewing was established with a web browser secured with Secure Socket Layer encryption.

### Expert visual analysis of samples

The expert assessment of physical slides was performed at the local pathology laboratory (Coast Provincial General Hospital, Mombasa, Kenya) with light-microscopy, performed by a trained pathologist (NM). Slides were initially classified as adequate or inadequate for examination. A slide was classified as inadequate for evaluation if the quality of the smear was unsatisfactory, it contained insufficient cellular material, lacked endocervical cells, had excess blood, mucus or other debris, or presented with high levels of inflammatory cells (covering >90% of cellular details). Physical slides classified as inadequate, which had already been digitized, were excluded as ineligible also from the validation series of digitized slides (n = 29). Slides which were adequate for analysis were reviewed by the pathologist, and results entered into the report form according to the Bethesda classification system.^34^ The acquired report form contained information about sample quality (satisfactory/unsatisfactory), evidence of infection or inflammation and detected atypia. For the analyses in this study, slides with findings recorded in the cytological report as low-grade squamous intraepithelial atypia (LSIL) or higher (i.e. ASC-H or HSIL, or higher) were included as slides with significant cervical-cell atypia. The expert assessment of the digital slides was performed by remotely located, independent experts. For this process, all slides in the validation series were initially screened by a cytotechnologist (KK) with experience in cervical-cytology screening, and slides with detected cellular atypia were reviewed by a pathologist with experience in Pap-smear analysis (LK). In accordance with generally accepted quality-control guidelines for cervical-cytology screening,^35^ 10% of slides that were assessed as negative in the initial cytological screening were randomly selected and submitted for re-evaluation by the pathologist. In this subset, no additional slides with significant atypia were detected. The samples were reviewed independently by the pathologist without access to the final cytodiagnoses made by the other pathologist or access to results from the final DLS-based analysis of samples. Visual analysis of slides was performed on a laptop computer with an LCD display using an internet browser to access and navigate the slides on the slide-management platform.

### Statistical analysis

General-purpose statistical software (Stata 15.1, Stata, College Station, TX, USA) was used for analysis of the results. The web-based REDCap application was used for collection and storage of patient data. Prior to analysis, the data were pseudonymized and exported into a standardized spreadsheet table (Microsoft Excel, Microsoft, Redmond, WA, USA). Statistical-power calculations were performed with a sample-size formula,^36^ assuming a prevalence (Pr) of 8% (± 2%) for significant atypia in the study population^37^ with α = 0.05 (and correspondingly Z1-α/2 = 1.96), and a precision parameter (ε) of 0.10, to determine whether sensitivity (SN) and specificity (SP) were comparable to the study ground truth.

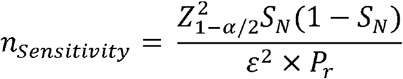

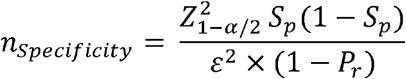

These calculations indicated a required target sample-size of 304 with the assumed prevalence of atypical samples. We estimated that 500 to 700 slides would be feasible to collect during the planned sample-acquisition period, given the frequency of visits for eligible patients at the clinic, and decided on a target number of 700 slides to yield 350 slides for training and 350 slides for validation. All statistical tests were two-sided unless otherwise stated. Diagnostic accuracy was evaluated in terms of sensitivity for the detection of abnormal slides and calculated as the percentage of true positives divided by (true positives plus false negatives). Specificity was calculated as the percentage of true negatives divided by (true negatives plus false positives). Evaluation of the performance of the algorithm was performed by calculating the AUC after plotting the measured true-positive rate (sensitivity) versus the false-positive rate (1 - specificity) for different thresholds of slide-level positivity. Statistical estimates of diagnostic accuracy are reported with 95% confidence intervals (95% CIs).

### Ethical statement

Approval for the current study was issued by the Ethical Review Committee at the National Commission for Science, Technology and Innovation (Pwani University, Nacosti, Kenya) (No. ERC/PU-STAFF/005/2018). The study was also approved by the Helsinki Biobank (No. 359/2017).

### Patient and public involvement statement

Patients and members of the public were involved at several stages of the project, including the design, management and execution of the study. When planning the study, we consulted with local co-investigators to identify actual needs of the local patients attending the clinic. Prior to study initiation, healthcare staff (community health volunteers and traditional midwives) provided information about the study to both eligible patients and the general public. During the study, personnel at the clinic (so-called mentor mothers and linkage officers) were in regular contact with the patients both at the clinic and outside the clinic, when disseminating results from the study. Patients were provided with information about the study purpose, methodology, procedures, and expected outcomes and were allowed to provide input and ask questions at all stages. For the ethical committee, we explicitly stated our goal to make the technologies here available locally, to ultimately benefit the general community. For this, we aim to disseminate the main results to the health authorities and focal people, including trial participants (patients attending the clinic), and will consult with our local collaborators to find an appropriate method to achieve this.

## Results

### Development of a deep-learning system for detection of cervical-cell atypia

To develop a deep-learning system for the detection of cervical-cell atypia in the digitized Pap smears, we used a commercially available machine-learning and image-analysis platform (Aiforia Create, Aiforia Technologies, Helsinki, Finland). Using this platform, we trained a machine-learning model based on deep convolutional neural networks, to detect LSILs and HSILs (or higher-grade lesions) in the Pap smear digital whole slides. The samples series was split with a 50-50 distribution of the target number of samples into the training series (*n* = 350), used for training and tuning of the model, and external validation series (*n* = 390). Individual digitized slides measured approximately 100,000 × 50,000 pixels. Training was performed by a researcher (OH), assisted by a cytotechnologist (KK) specialized in cervical-cytology screening, using manually defined representative regions of the digitized slides of the training series (Figure 2).

**Figure 2.**
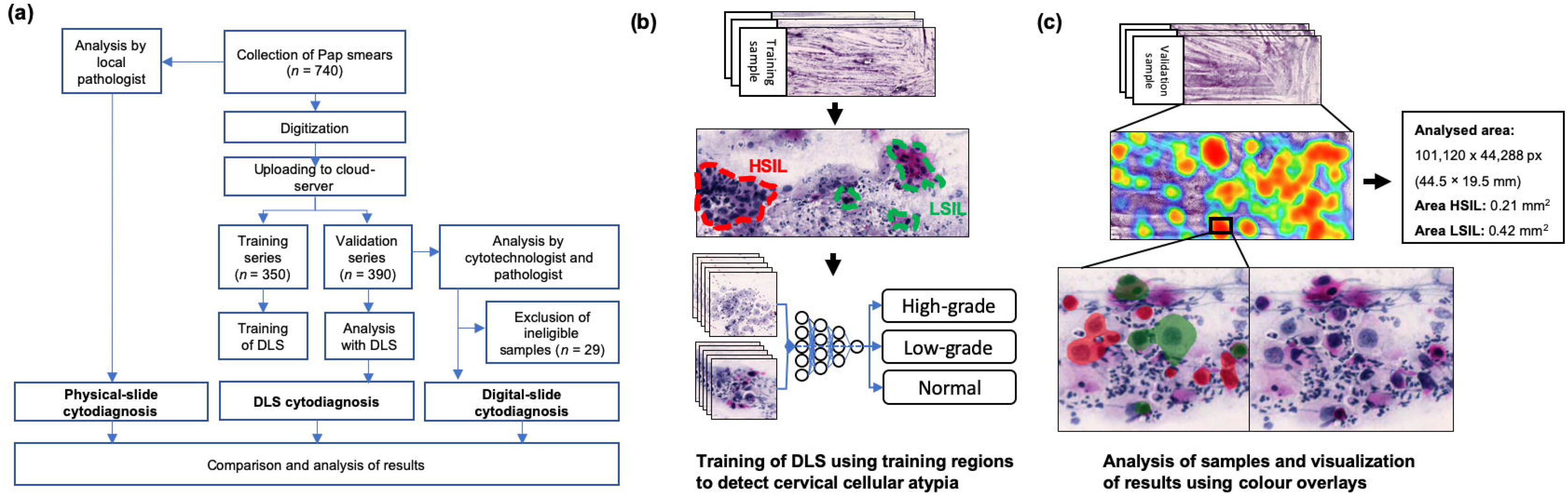
Overview of sample processing and algorithm training and validation. (a) Flowchart illustrating the sample-processing workflow, showing stages from the collection of samples to the analysis of digital images and physical slides. (b) Schematic view of the annotation process used for creation of the digital-slide data for training of the deep-learning system (DLS). (c) Validation analysis of a digitized image of a whole slide (Pap smear) with the DLS, showing calculations of areas of atypia, with locations of atypia in a heatmap of the digital slide, and identification of individual cells, with colour overlays (red for high-grade atypia and green for low-grade atypia).

Regions (*n* = 16,133, with cross-sections of ~25-100 μm) were selected visually, and included areas of both normal cervical cellular morphology and various degrees of atypia; visible atypia (low-grade and high-grade) was manually annotated.

Training of the DLS used 30,000 iterations (training epochs), with a pre-determined feature size (field-of-view) of 30 μm. To optimize the generalizability of the deep-learning algorithm, the training data were augmented with the following image perturbations: variation in scale (± 10%), aspect ratio (± 10%), shear distortion (± 10%), luminance (± 10%), contrast (± 10%), white balance (± 10%) and variation in image compression quality (40-60%). DLS analyses were performed on entire digitized slides, and a slide-level operating threshold for the total area of detected atypia was decided on the basis of the training data, to determine whether slides were classified as atypical, decided on based on best performance in the training data set. Analysis time for one whole-slide image with the trained model was ~30 s. The image analysis gave the total area per slide of LSILs and HSILs (or higher-grade lesions). For the slide-level classification by the DLS, slides with both detected low- and high-grade lesions where classified as high-grade.

### Detection of cervical-cell atypia in digital Pap smears with the DLS

Following the training of the DLS with the training slides (*n* = 350), we analysed the slides in the validation series with the DLS to detect cervical-cell atypia, and compared the results to the ground-truth assessment of digital and physical slides. After exclusion of inadequate slides (29; 7%), 361 slides remained in the validation series. Expert assessment of digitized slides by the cytotechnologist and pathologist revealed 19 slides (5%) with low-grade atypia, 28 slides (8%) with high-grade atypia and 314 slides (87%) that were negative for significant squamous cell atypia (defined as atypical squamous cells of undetermined significance (ASC-US) or lower).

Compared with these expert assessments, for the detection of slides with general atypia (low-grade and/or high-grade atypia), the DLS achieved an area under the receiver operating characteristic curve (AUC) of 0.94 and, at the operating threshold, sensitivity of 95.7% (95% CI 85.5–99.5%) and specificity of 84.7% (95% CI 80.2–88.5%) (Figure 3). The AUC for detection of slides containing HSILs or higher-grade lesions was 0.93, with sensitivity of 85.7% (95% CI 67.3 - 96.0%) and specificity of 98.5% (95% CI 96.5–99.5%) at the chosen threshold. For the detection of slides containing only LSILs, the AUC was 0.86, with sensitivity of 84.2% (95% CI 60.4–96.6%) and specificity of 86.0% (95% CI 81.8–89.5%) at the selected threshold (Table 1). In these analyses, discrepancies in the type of atypia (such as LSIL-only slides that were classified by DLS as high-grade, or vice versa) were considered to be of equal statistical value to slides with atypia that were classified by DLS as negative. Overall, the DLS classified 266 slides (74%) as negative, 61 slides (17%) as positive for low-grade atypia and 34 slides (9%) as positive for high-grade atypia. Compared to the expert assessments, two slides with low-grade atypia were falsely classified as negative by the DLS (<1%), but no slides with high-grade atypia were falsely classified as negative by the DLS, although four slides (1%) with high-grade atypia were classified as low-grade atypia by the DLS (Table 1). The negative predictive value (NPV) for the DLS was high for general atypia (266/268 = 99.3%; 95% CI 97.3–99.9%), low-grade atypia (294/297 = 99.0%; 95% CI 97.1–99.8%) and high-grade atypia (328/332 = 98.8%; 95% CI 96.9–99.7%). Examples of comparisons between DLS assessments of atypia and expert assessments at the cellular level are shown in Figure 3. In summary, compared to the expert assessment of the digitized slides, the DLS demonstrated high sensitivity for the detection of cervical-cell atypia in general, with higher diagnostic accuracy for high-grade than for low-grade atypia. The total number of false negatives was low, and NPV high for atypical slides.

**Figure 3.**
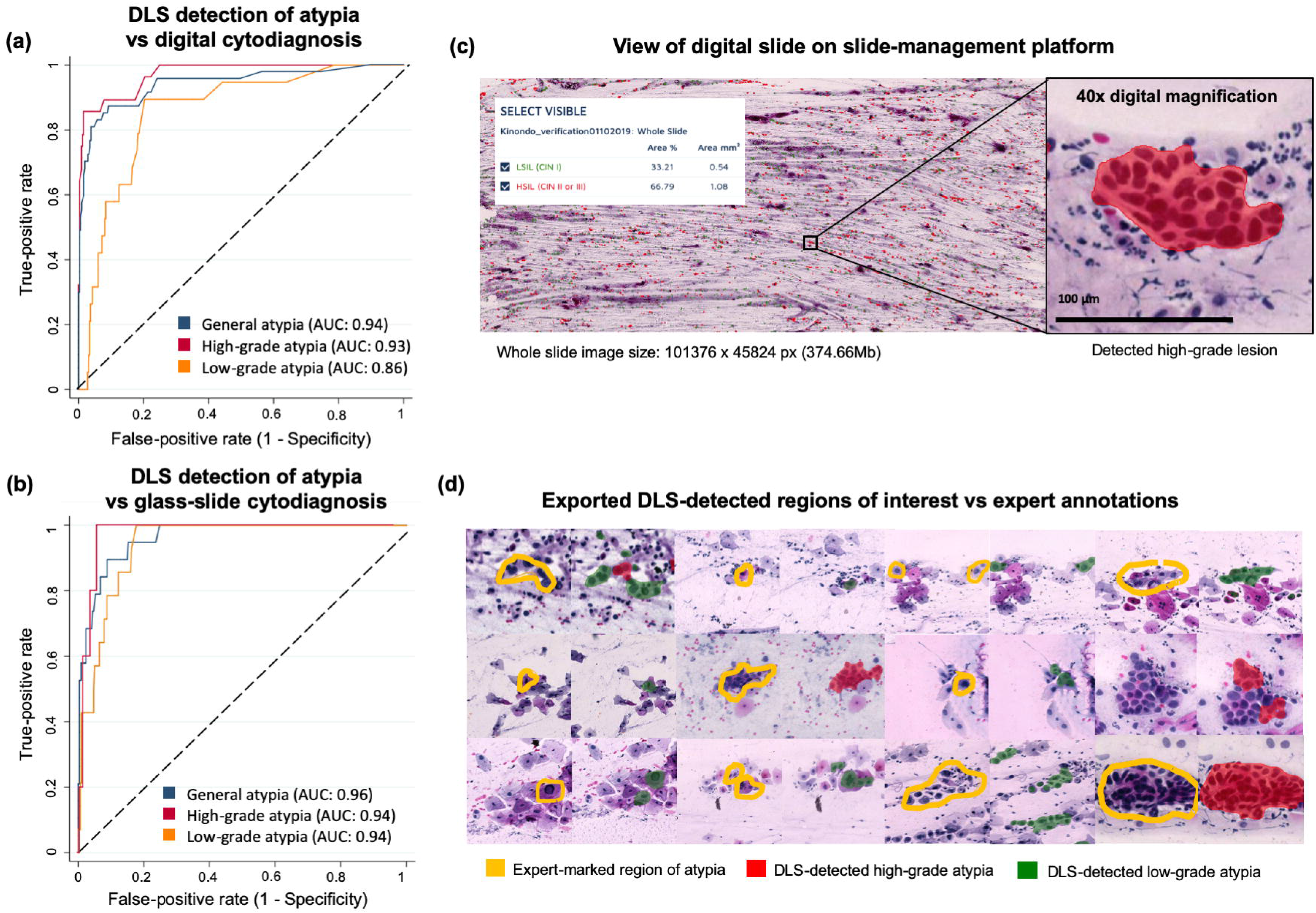
Detection of atypia in cervical smears by automated deep-learning system (DLS) and by manual assessment. **(a)** and **(b)** Areas under the receiver operating characteristic (ROC) curves (AUCs) for the detection of general atypia, high-grade atypia and low-grade atypia with the DLS compared with manual assessment of **(a)** digital slides by a cytotechnologist and a pathologist, and **(b)** physical slides by a local pathologist. ROC curves were calculated for a range of operating thresholds for the DLS, and the diamond marker on each ROC curve corresponds to thresholds used for the model for each assessment. **(c)** View of a digitized sample on the cloud-based slide-management platform, with a magnified view of a detected atypical cellular cluster at 40x digital magnification. **(d)** Examples of atypical cells marked by the experts in the digitized slides (yellow), and the corresponding regions extracted from the DLS results, with cells assessed as high-grade atypia coloured in red and low-grade atypia coloured in green.

**Table 1.**
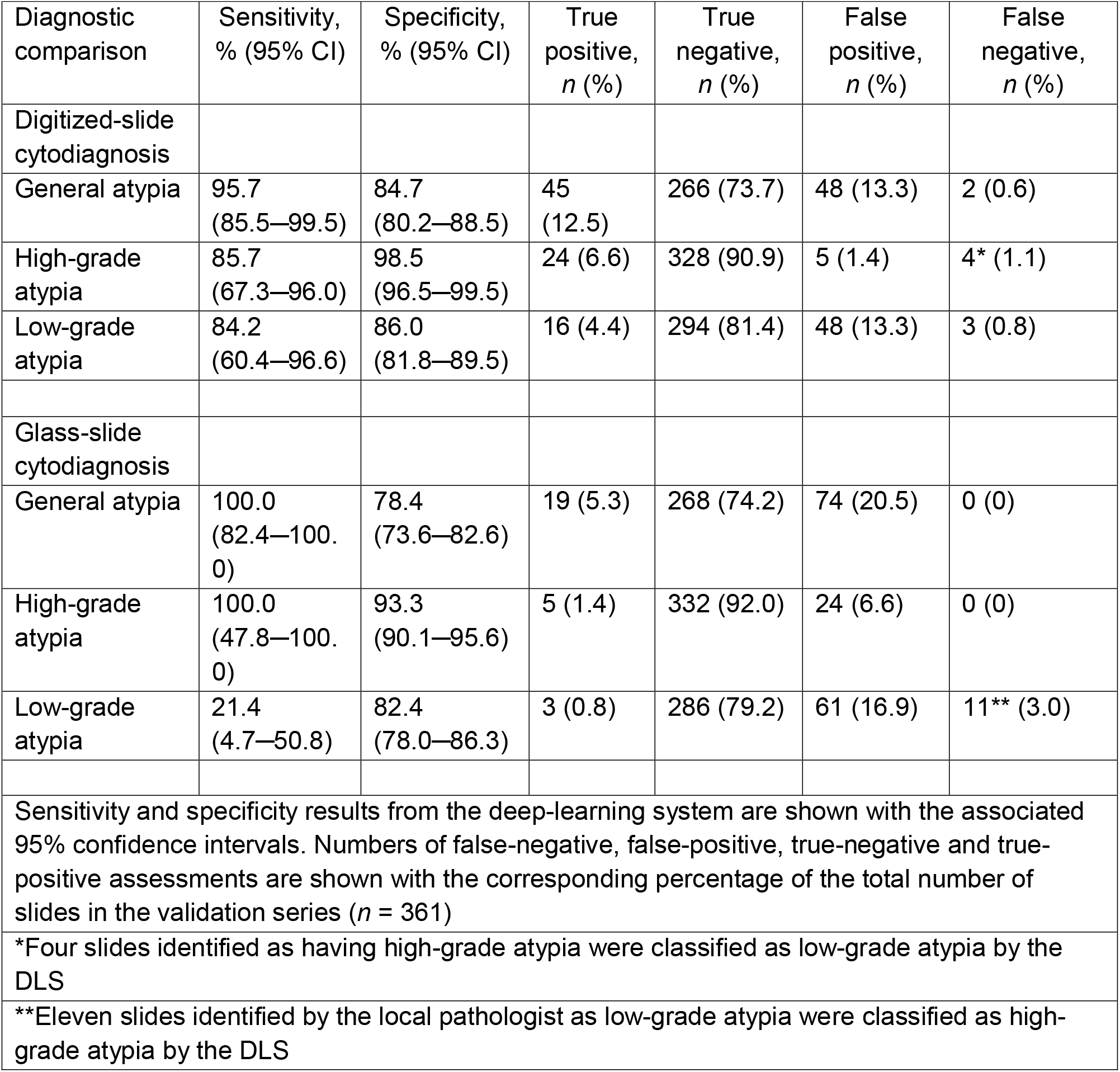
Detection of cervical-cell atypia with the deep-learning system (DLS) in digitized Pap smears, compared with expert-assessments of digitized and physical slides.

### Comparison of DLS results to the pathologist glass-slide cytodiagnosis

Next, we evaluated the performance of the DLS for detection of cervical-cell atypia, as compared to the assessment of physical slides, performed by a local pathologist. To do this, we compared the results obtained by the DLS with those produced by the local pathology laboratory for physical-slide cytodiagnosis. In the cytological report of the 361 physical slides in the validation subset, 342 (95%) were classified as negative for significant squamous-cell atypia, 14 (4%) were classified as positive for low-grade atypia and five (1%) were classified as positive for high-grade atypia. With reference to these results, the DLS achieved high sensitivity for general atypia (100%; 95% CI 82.4-100%) and for high-grade atypia (100%; 95% CI 47.8-100%), with corresponding specificities of 78.4% (95% CI 73.6–82.4%) and 93.3% (95% CI 90.1–95.6%), respectively (Table 1). Specificity was moderate for low-grade atypia (82.4%; 95% CI 78.0–86.3) but sensitivity was lower (21.4%; 95% CI 4.7–50.8) as 11 of the 14 slides that were classified as low-grade atypia in the cytological report from the pathology laboratory were classified as high-grade atypia by the DLS. The NPV was high for general atypia (266/266 = 100%; 95% CI 98.6–100.0%), high-grade atypia (332/332 = 100%; 95% CI 98.9–100.0%) and low-grade atypia (286/297 = 96.3%; 95% CI 93.5–98.1%) (Table 1). The DLS achieved high AUCs for detection of general atypia (0.96), high-grade atypia (0.94) and low-grade atypia (0.94) (Figure 3). Compared to the physical-slide cytodiagnosis, no atypical slides were falsely classified as negative by the DLS. In conclusion, we observed a high sensitivity for the DLS for detection of slides classified as atypical by the local pathologist, but the DLS showed a lower threshold for classifying slides as high-grade, resulting in a majority of low-grade slides being classified as high-grade by the DLS.

## Discussion

In this study, we implemented a POC digital diagnostics system at a peripheral clinic in Kenya and evaluated it for the analysis of Pap smears. The DLS achieved high diagnostic accuracy for the detection of slides with cervical squamous cell atypia, with AUCs of 0.94–0.96 and sensitivities of 96-100%, compared to the visual interpretation of digitized and physical slides. With the visual assessment of digitized slides as reference, the number of false-negative assessments by the DLS was low, with two low-grade and no high-grade slides being incorrectly classified as negative (although four high-grade slides were falsely classified as low-grade). Compared to the visual analysis of the physical slides by the local pathologist, the DLS sensitivity was high for general atypia (100%) and high-grade atypia (100%) but was low for low-grade atypia (21%), as 11 of 14 physical slides that were assessed as low grade were classified as high grade by the DLS. The visual interpretation of Pap smears is known to be subjective, especially when assessing low-grade findings,^10,38^ and accordingly, we observed variation between the expert assessments of slides, with a lower threshold for the classification of findings as high grade by the pathologist who assessed the digitized slides. The DLS was trained with assistance from one of the experts who analysed the digitized training slides, which possibly explains why the DLS classification resembled the expert assessment of digitized slides. Notably, however, none of the slides that were classified as negative by the DLS were classified as atypical in the cytodiagnosis of the physical slides.

Previous studies have reported encouraging results with the deep learning-based analysis of smaller cropped images from Pap smears^26,27,29,39^ that were digitized with conventional slide scanners, but clinical application requires the examination of substantially larger sample areas.^28^ In this study, we used routine samples collected in the clinic, and correspondingly, the whole-slide images were magnitudes larger than those previously analysed, measuring on average 100,387 × 47,560 pixels; thus, the total number of pixels analysed corresponded to approximately twice the number in the entire ImageNet database at commonly used resolutions.^40^ Pap smears may contain very limited numbers of isolated atypical cells, and robust algorithms are necessary to reliably detect such cells in these large and complex samples. In this study, we investigated the use of a DLS as a potential screening tool with a relatively low threshold for the classification of slides as atypical, to ensure high sensitivity at the potential expense of specificity; this method resulted in relatively high rates of false positives for low-grade atypical slides (13-17%). However, as this type of algorithm can operate using multiple configurations, sensitivity and specificity could be adjusted to match clinical requirements, with high sensitivity for screening purposes or higher specificity for confirmatory diagnostics. Importantly, our findings demonstrate how a front-line diagnostic system based on POC digital microscopy with the deep learning-based analysis of whole microscopy slides can be deployed in rural clinical settings. To our knowledge, no other study has evaluated this technology using whole slides that have been collected, stained, digitized and uploaded using a mobile data network in a similar setting. Overall, we achieved high NPVs for the detection of atypical slides, suggesting that this technique may be useful for screening purposes. For this application, clinical implementation could reduce sample analysis workloads to allow clinicians to focus on verifying potentially abnormal slides and could exclude the majority (~70%) of slides while retaining high sensitivity for atypical slides. By combining this technology with primary POC molecular testing for HPV^8,14^, the number of slides that needs to be analysed could be reduced even further, which would be essential in low-resource areas where the number of practising pathologists is low and the cervical cancer incidence is rising.^4,41^ By using methods such as self-sampling for both molecular and cytology-based testing,^42^ the dissemination of tests to large populations could be feasible. The importance of fast, accurate and reliable diagnostics is especially important for patients with risk factors, such as HIV positivity, who are not only less suitable for screening with only HPV tests but also are at a higher risk for other diseases, such as sexually transmitted infections,^43^ neglected tropical parasites^44^ and other malignancies.^45^ Notably, as these conditions are currently diagnosed with microscopy, the technology described here is likely to also be applicable for diagnostics of these diseases,^2,46,47^ thus creating opportunities for integrated disease control.

As this is an early study, it had some limitations. The performance of the DLS was benchmarked against two independent experts for the assessment of cytological samples, but for the results to be directly comparable to other screening modalities, the ideal reference standard would be cervical biopsies with histologically confirmed precancers, which were not available here. Further, even though the total number of slides collected was relatively large, the prevalence of slides with significant atypia was limited. Although these results are promising, increasing the amount of training data would likely improve the performance of the DLS, and would be required before confirmatory diagnostic applications. To achieve this, experts who would eventually be using the system should ideally be involved during the training process of the DLS to ensure consistent and reliable annotations of training data. Moreover, as this study was a single-centre study, the results might differ if the sample acquisition and preparation procedures are altered, and further work is needed to prospectively validate these results. As the technology here provides a platform for digital microscopy at the POC, which is likely to also be useful for diagnostics of other conditions, future studies are also warranted to evaluate other potential applications.

## Conclusion

We have developed a novel system for deep learning-based digital microscopy at the POC, which was used for the analysis of cervical smears in cervical cancer screening. The detection of squamous cell atypia with the technology is feasible, with high sensitivity for slides demonstrating atypia, and particularly for slides showing high-grade atypia. The clinical utilization of this technology could reduce the sample analysis workload for microscopists and provide a platform for general-purpose digital pathology, which is implementable in rural areas. As such, the technology here could create novel opportunities to facilitate the diagnostics of a variety of diseases that are still underdiagnosed, especially in low-resource settings.

## Data Availability

The software used to develop the deep learning-algorithm in this study is commercially available at: https://www.aiforia.com/aiforia-create/, and the parameters and configurations required for replications of results are described in the manuscript. We aim to make the final algorithm publicly available for analysis of digital samples three (3) months following publication of the article. The model will be made available as an application program interface (API). Further requests for sharing of de-identified data (digitized samples) will be considered from researchers abiding the following principles: data will be securely stored with appropriate documentation and not disposed into publicly accessible domains or otherwise shared without explicit permission from the authors, data is only used with the aim to generate data for the public good. Applications are subjected to review by the Data Access Committee at Institute for Molecular Medicine Finland (FIMM), University of Helsinki, Helsinki, Finland)

## Author contributions

Conceived and designed the experiments: OH, NL, HK, VD, JL. Collected data: NM, JM, FK, ST, MM. Performed the experiments: OH, NM, FK, MM, ML, JL. Analysed the data: OH, NM, FK, MM, LK Contributed reagents/materials/analysis tools: NL, HK, JM, FK, ST, MM, ML, JL. Wrote the manuscript: OH, NL, JL Contributed to the manuscript revision: OH, NL, FK, MM, LK, ML, VD, JL. Supervised the project: JL, HK. All authors contributed to the editing, review and final approval of the manuscript. The corresponding author confirms that he had full access to all the data in the study and had final responsibility for the decision to submit for publication

## Acknowledgments

The Erling-Persson Family Foundation funded this study. In addition, this work has been supported by the Swedish Research Council, Sigrid Jusélius Foundation, Finska Läkaresällskapet and Medicinska Understödsföreningen Liv och Hälsa rf. In addition, this project has received funding from K. Albin Johanssons stiftelse, Stiftelsen Dorothea Olivia, Karl Walter och Jarl Walter Perkléns Minne, Wilhelm och Elsa Stockmanns stiftelse and Biomedicum Foundation.

We thank the FIMM Digital Microscopy and Molecular Pathology Unit supported by Helsinki University and Biocenter Finland for outstanding assistance. We would like to greatly acknowledge the work with data collection by Caroline Jeptoo and Priscillah Kinyanjui, the IT-support from Hakan Küçükel, the assistance with analysis of samples by Kaija Knuuti and assistance from Sanni Ruotsalainen with statistical analyses. Furthermore, we would like to acknowledge Pekka Nieminen and Stig Nordling for consultations during the study planning.

## Data availability

All data required to evaluate the conclusions in the article are included in the manuscript and/or the Supplementary Materials. Additional data is available on request from the authors, and applications handled by the Data Access Committee at Institute for Molecular Medicine Finland (FIMM), University of Helsinki, Helsinki, Finland)

## Data availability

The software used to develop the deep learning-algorithm in this study is commercially available at: https://www.aiforia.com/aiforia-create/, and the parameters and configurations required for replications of results are described in the manuscript. We aim to make the final algorithm publicly available for analysis of digital samples three (3) months following publication of the article. The model will be made available as an application program interface (API). Further requests for sharing of de-identified data (digitized samples) will be considered from researchers abiding the following principles: data will be securely stored with appropriate documentation and not disposed into publicly accessible domains or otherwise shared without explicit permission from the authors, data is only used with the aim to generate data for the public good.

Applications are subjected to review by the Data Access Committee at Institute for Molecular Medicine Finland (FIMM), University of Helsinki, Helsinki, Finland)

## Supplementary material

**Supplementary Figure 1**. CONSORT-style flowchart illustrating flow of samples in study. (Separate file)

**Supplementary Table 1**. Epidemiological information and patient cohort characteristics

